# An accurate and rapid Real-time PCR approach for human Monkeypox virus diagnosis

**DOI:** 10.1101/2022.06.23.22276033

**Authors:** Tony Wawina-Bokalanga, Nikola Sklenovska, Bert Vanmechelen, Mandy Bloemen, Valentijn Vergote, Lies Laenen, Emmanuel André, Marc Van Ranst, Jean-Jacques Muyembe-Tamfum, Piet Maes

## Abstract

**Background:** Human monkeypox (MPX) is an emerging zoonotic disease caused by monkeypox virus (MPXV), which is endemic to Western and Central Africa. Despite being identified in captive monkeys in 1958, and in humans from the Democratic Republic of Congo (DRC) in 1970, a MPXV diagnostic test is not routinely implemented in diagnostic laboratories. Currently, a multi-country outbreak of MPXV has been ongoing in several European and American countries since May 2022.

**Methods:** To develop this real-time PCR assay, five MPXV positive DNA samples were tested using MPXV-specific primers. The qPCR amplification was conducted on two different qPCR devices. We also tested samples, collected in May 2022, from pox-like lesions of MPXV-suspected patients in Belgium.

**Results:** The performance of our real-time PCR was proved with five MPXV positive DNA from DRC. In addition, five out of ten clinical samples, from the current MPXV-outbreak, tested positive with Ct values ranging from 20.6 to 34.9.

**Conclusions:** In this study, we present an accurate and rapid real-time PCR approach, based on conserved regions of both MPXV clades, that can be used to test clinical samples from MPXV-suspected persons. Our real-time qPCR assay can be used for the 2022 multi-country outbreak of human MPXV.

**Summary:** An accurate and fast real-time PCR assay is indispensable to diagnose cases of monkeypox during the 2022 multi-country outbreak.

## INTRODUCTION

Monkeypox virus (MPXV) is a double-stranded DNA virus that belongs to the genus *Orthopoxvirus* within the family *Poxviridae*, subfamily *Chordopoxvirinae*. This genus consists of 12 different species of which four contain viruses that are pathogenic to human: *Monkeypox virus, Vaccinia virus, Cowpox virus*, and *Variola virus*. The latter is the causative agent of smallpox, a disease which has been globally eradicated since 1977 [1, 2].

Following the first reported cases of human monkeypox infections in 1970 in the Democratic Republic of Congo (DRC, formerly Zaïre), other human MPXV outbreaks have been reported in Central and West Africa [3]. In 2003, the United States of America (USA) experienced a human MPXV outbreak in multiple states, the first one in the Western Hemisphere [4, 5]. In addition, monkeypox infections have been reported in travelers from Nigeria to Israel (September 2018), to the United Kingdom (September 2018, December 2019, and May 2021), to Singapore (May 2019), and to the USA (July and November 2021) [6-8].

From May 2022, a multi-country outbreak of MPXV has been ongoing in several countries, primarily in Europe and the Americas. The first cases of this outbreak were mainly but not exclusively identified amongst men who have sex with men seeking treatment in primary care and sexual health clinics [9].

Despite being identified in captive monkeys already in 1958, and in humans from DRC in 1970, there is no certified MPXV in-vitro diagnostic (IVD) kit that can be used for human MPXV detection. Furthermore, the lack of certified MPXV IVD kits could negatively impact the surveillance of MPXV transmission in the community. Therefore, different PCR methods mainly based on real-time PCR have been proposed and used in monkeypox endemic regions [10-13].

Human monkeypox infection is endemic in West and Central African countries, where MPXV remains the most pathogenic poxvirus. The case fatality rate in Africa varies between 1 and 11% depending on the MPXV clade, the surveillance system and health care infrastructures [14, 15].

MPXV-infection is characterized by a 1-to 4-day prodromal period (with fever and malaise) before development of skin rashes with indurated and umbilicated lesions, starting on the head or face and progressing to the limbs and trunk (Table 1) [16, 17].

**Table 1.**
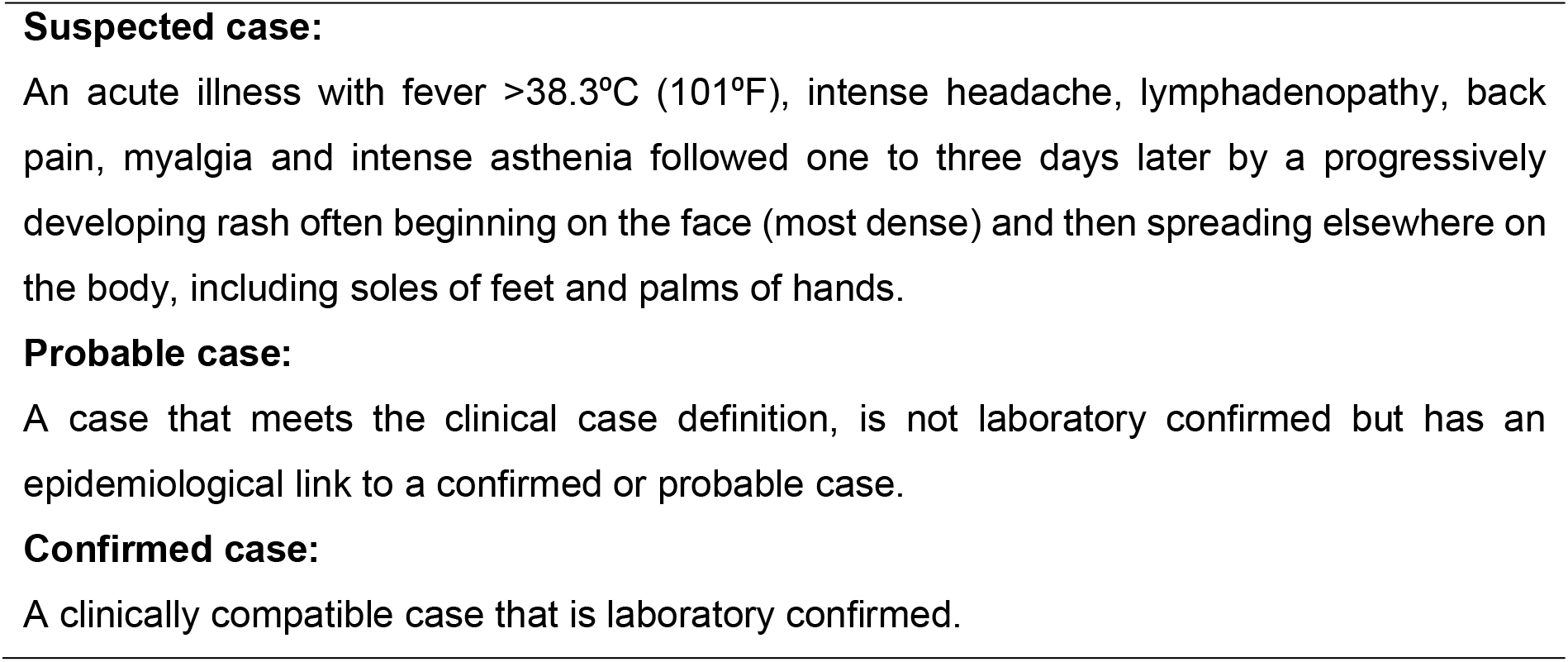
WHO case definition^[18]^

Furthermore, the rising number of MPXV-infected people over the last years can be associated with cessation of routine vaccination against smallpox, which increased human vulnerability to MPXV [19].

Prior to the current outbreak, our laboratory had developed an accurate and rapid real-time PCR approach that can be used for human MPXV diagnosis. In this report, we show the validation of this approach and illustrate its employability as a diagnostic tool for the current MPXV outbreak.

## METHODS

### Sample collection

Five MPXV positive DNA samples collected in 2016 were kindly provided by the Institut National de Recherche Biomédicale (INRB), Kinshasa (DRC). These DNA samples were extracted locally at INRB from skin lesions of MPXV-infected patients.

Viral DNA from the pox-like lesions of MPXV-suspected patients in Belgium, collected in May 2022, was extracted using the MagMax viral pathogen II nucleic acid isolation kit for KingFisher™Flex system (ThermoFisher).

### Primer and probe design

Selection of MPXV-specific primers and hybridization probes was performed based on the sequence of a selection of MPXV genomes from the Central Africa clade, which were retrieved from the GenBank database and multiple-aligned using MUSCLE in MEGA v7.0 [20].

Open reading frame O2L (662 bp in length), one of the conserved regions of MPXV genomes, was used as a template for designing the specific primers and probes through the Integrated DNA Technologies, PrimerQuest™tool [21].

Oligonucleotide primers and corresponding hybridization probes were checked for self-complementarity and optimal melting temperature using the online program Oligo Calc [22]. The MPXV-specific probe contained the 6-FAM/ZEN-IBFQ dye/quencher pair, which has a maximum emission wavelength of 518 nm (Table 2).

**Table 2.**
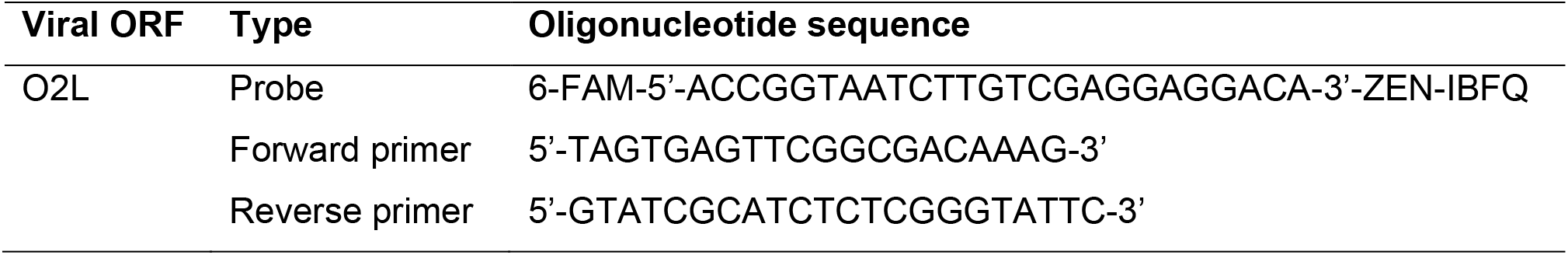
Validated MPXV-specific primers and probe

### Real-time PCR amplification

PCR amplification was conducted on Mic qPCR (Bio Molecular systems) and AB™7500 Fast Real-Time PCR system (Applied Biosystems, USA) devices. Reactions were performed using a 20 µl reaction mixture containing 5 µl 4x TaqMan™Fast Virus 1-Step Master Mix, 900 nM of each forward and reverse primers, 250 nM probe, and 5 µl DNA template. Each qPCR reaction was performed in duplicate. PCR reactions were performed under the following conditions: 20 sec at 95°C and 45 cycles of 3 sec at 95°C and 30 sec at 60°C (Fast Mode).

### Confirmation through Sanger sequencing

The PCR products were purified using the ExoSAP-IT kit (Applied Biosystems, USA). Cycle sequencing of the purified PCR products was performed using BigDye Terminator chemistry (Applied Biosystems, USA). Sequencing reactions were purified with a rapid in-house method using 1:10 volume sodium acetate (3M, pH 5,5) mixed with 70% ethanol. The purified sequencing products were run on ABI PRISM 3100 (Applied Biosystems, USA). Sequences were analyzed and corrected using SeqMan software, version 7.0.0. To confirm the specificity of our primers, the obtained sequences were compared to a MPXV reference genome using BLAST tool [23].

### Analysis of clinical samples from the ongoing human MPXV outbreak

Clinical samples (pox-like lesions) from MPXV-suspected patients were collected by physicians then transported safely to the laboratory in accordance with national and international requirements. Viral DNA was extracted using MagMax viral pathogen II nucleic acid isolation kit for KingFisher™Flex system (ThermoFisher).

PCR amplification was conducted on QuantStudio™7 Flex Real-Time PCR System. Reactions were performed using a 20 µl reaction mixture 5 µL TaqMan™Fast Virus 1-Step Master Mix (Applied Biosystems, Cat 4444434), 900 nM of each forward and reverse primers, 0,5 µL probe, 7,5 µL RNase free water and 5 µL DNA template. Each qPCR reaction was performed in duplicate under the PCR conditions mentioned previously (Figure 1).

**Figure 1.**
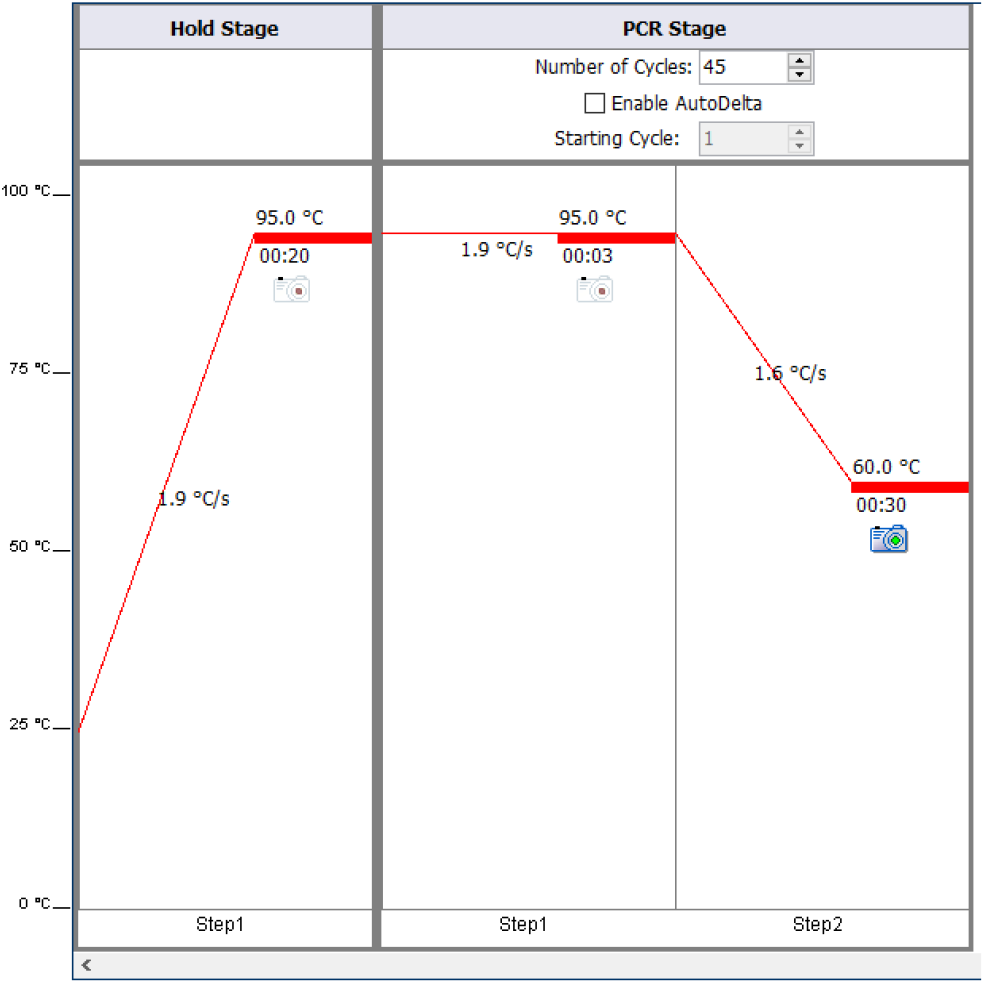
PCR conditions for MPXV diagnosis on QuantStudio™7 Flex RT PCR System

### Ethics

Ethical permission for the work conducted at KU Leuven on the viral DNA samples was approved by the KU Leuven ethics committee under reference no. S58836.

## RESULTS

### Real-time PCR assay on AB™7500 Fast Real-Time PCR system

All five MPXV DNA samples used for assay development were positive with a cycle threshold value (Ct) ranging from 15.3 to 24.6 (Figure 2).

**Figure 2.**
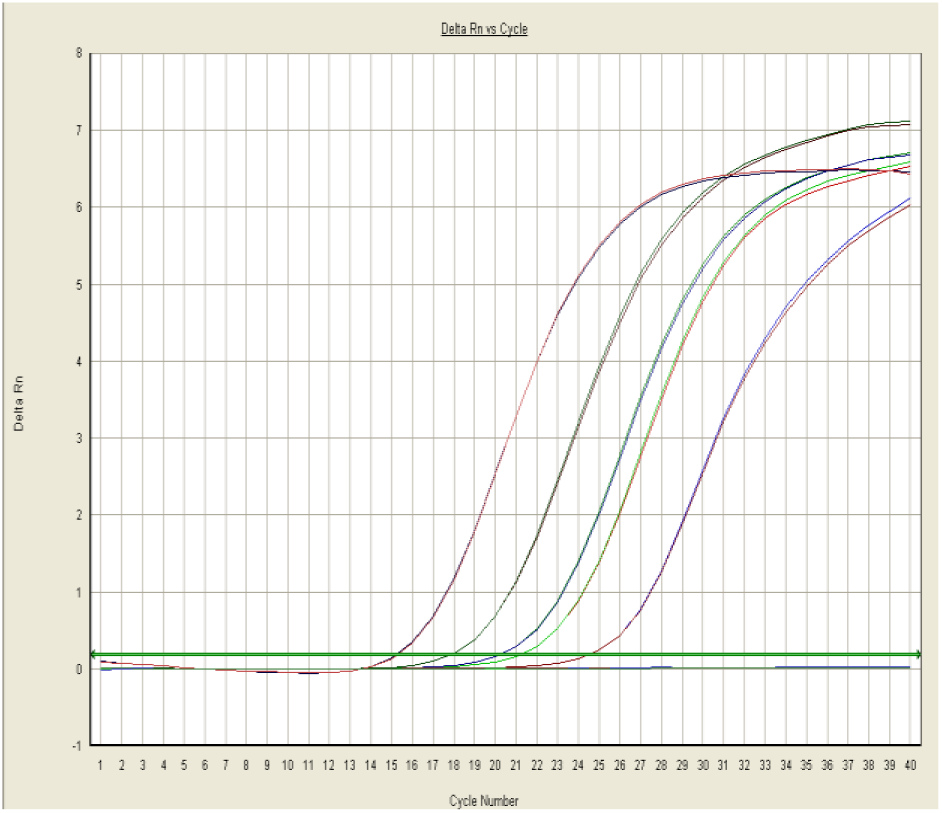
Δ Rn vs cycle representation of the real-time PCR performed on an AB™7500 Fast Real-Time PCR system. Data were obtained using the 7500 Fast System Software v1.3.1 with primers and hybridization probe calculated for MPXV. The detection signal of 6-FAM dye conjugated with MPXV-probe is 518 nm.

### PCR assay on Mic qPCR cycler

To assess the sensitivity of our RT-PCR assay, we performed 10-fold serial dilutions of each positive MPXV-DNA sample on a Mic qPCR cycler and the results were analyzed using the MicPCR Software v2.2.0. As seen on the AB™7500 Fast Real-Time PCR system, all dilutions from each positive MPXV-DNA sample were confirmed to be MPXV-positive with Ct values ranging from 15.3 to 22.3 (Figure 3).

**Figure 3.**
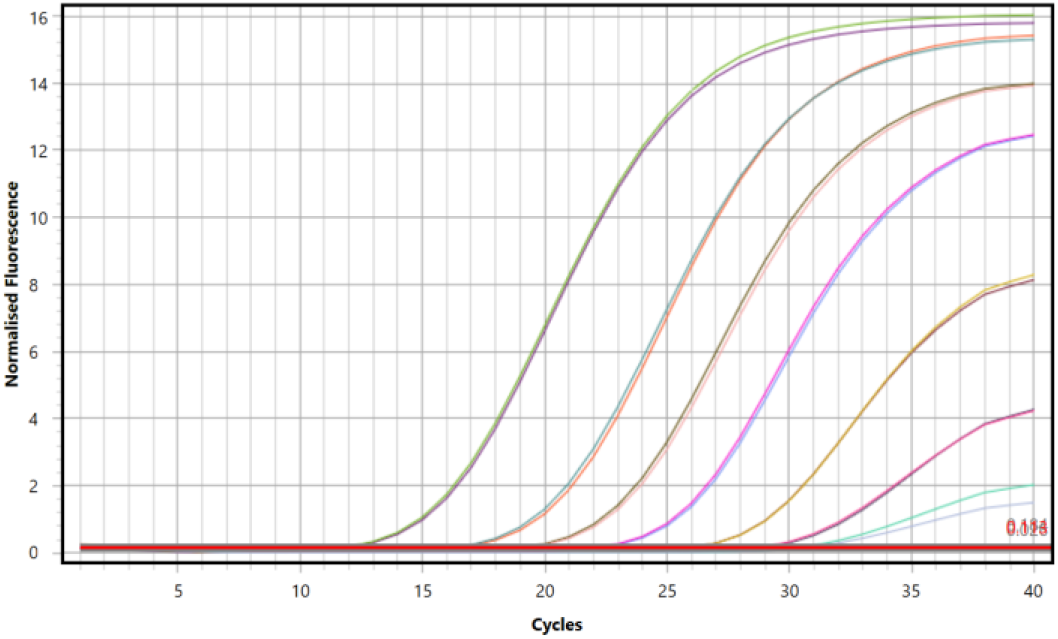
Δ Rn vs cycle representation of the qPCR performed on the MIC cycler. MPXV PCR amplification curves from successive 10-fold serial dilutions of a positive sample with Ct values ranging from 15.3 to 22.3.

### Validation of our primers and probe for the current outbreak strain

To confirm the specificity of our primers and probe, a BLAST search was performed against all available genomes of both MPXV clades, including sequences from the recent outbreak. The query coverage and maximum identity of our primers were 100% for both MPXV clades. However, a single mismatch (ACCGGTAATCTTGTCGA**<u>T</u>**GAGGACA) with genomes of the West African clade, including those from the current outbreak, was observed in the probe.

To validate whether the qPCR assay was still efficient, we conducted our qPCR on a 5-fold serial dilution series of a MPXV-suspected sample from the ongoing multi-country outbreak. A DNA sample from the second reported case of MPXV-infection in Belgium was used and all serial dilutions were confirmed MPXV positive, with Ct values ranging from 16.5 to 37.3 as shown in Figure 4.

**Figure 4.**
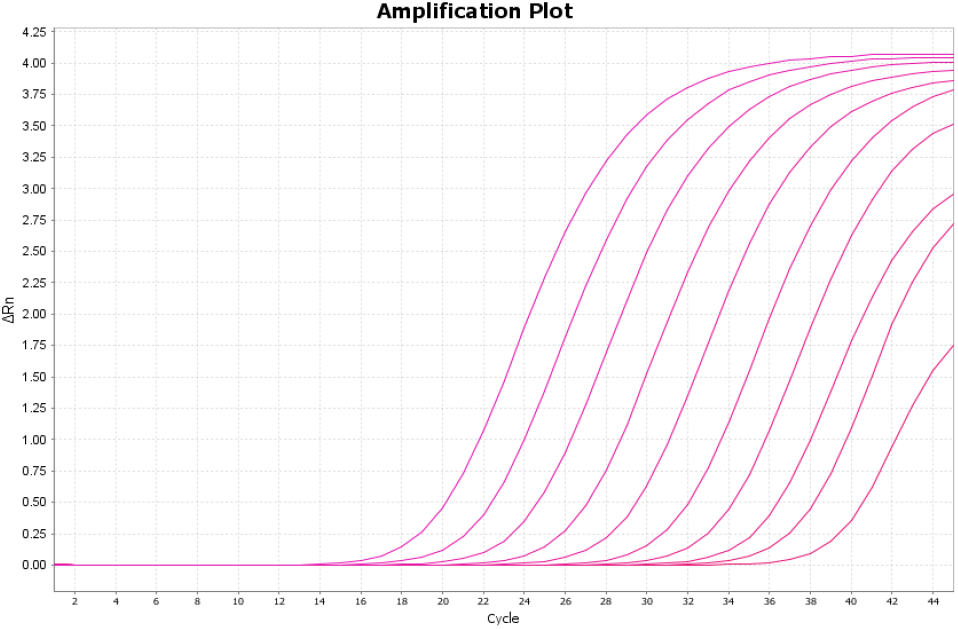
Δ Rn vs cycle representation of the qPCR performed on the QuantStudio™7 Flex Real-Time PCR System. MPXV PCR amplification curves from successive 5-fold serial dilutions of a positive sample with Ct values ranging from 16.5 to 37.3.

**Figure 5.**
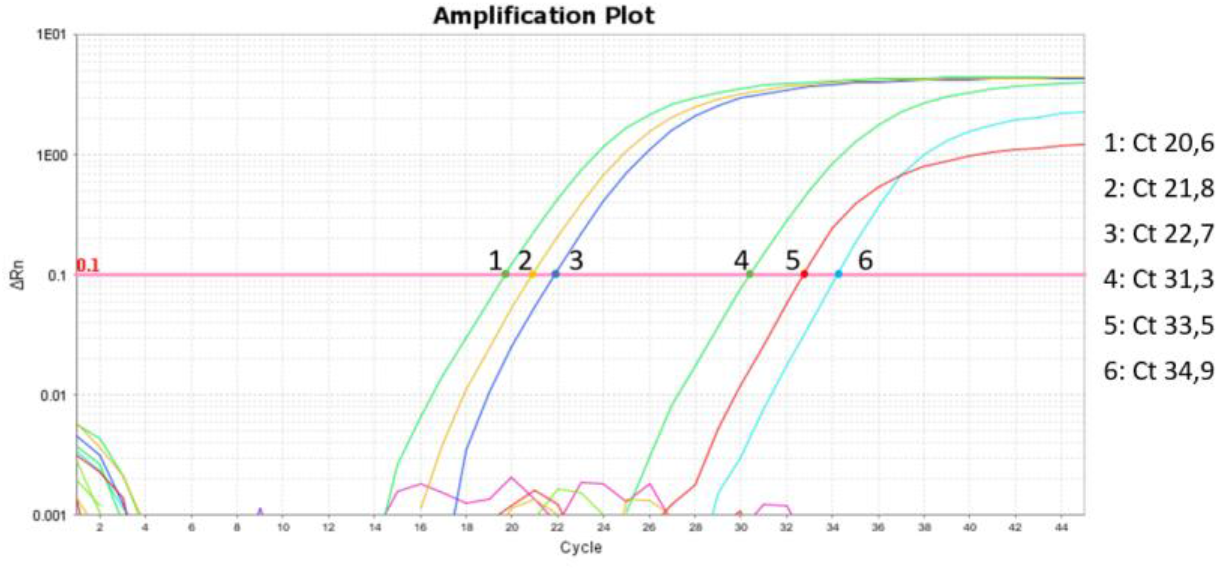
Δ Rn vs cycle representation of the qPCR performed on the QuantStudio™7 Flex Real-Time PCR System. PCR amplification curves of DNA samples from the ongoing human MPXV-outbreak, which started in May 2022. 1: Positive control, 2-6: Clinical DNA samples.

Following this initial sample, an additional ten MPXV-suspected samples were tested using our qPCR. Five out of ten clinical samples tested positive, with Ct values ranging from 20.6 to 34.9.

## CONCLUSION

In this study, we confirmed the usability of a previously in-house developed qPCR assay for the diagnosis of MPXV-infected samples. The initial assay validation was performed on both the Mic qPCR and AB™7500 Fast Real-Time PCR systems. The used primers are specific for all known MPXV genomes, and while the probe contains a single mismatch with the currently circulating strain, it can still reliably detect viruses of both the Central and West Africa MPXV clades, including the clade that is currently circulating in Europe.

In summary, this qPCR assay has been proven to be MPXV-specific, highly efficient and rapid. Furthermore, it can be used for the 2022 multi-country outbreak of MPXV.

## Data Availability

All data produced in the present study are available upon reasonable request to the authors

## Author Contributions

First draft preparation: **TWB**. Conceptualization and investigation: **TWB, NS, MB, VV, MVR, PM**. Writing-review and editing: **TWB, NS, BV, LL, EA, JJMT**, and **PM**. All authors have read and approved the final version of the manuscript.

## Acknowledgements

The authors thank the Institut National de Recherche Biomédicale (INRB), through Professor J.J Muyembe Tamfum, for providing the positive MPXV-DNA samples.

